# CSF proteomic profiling with amyloid/tau positivity identifies distinctive sex-different alteration of multiple proteins involved in Alzheimer’s disease

**DOI:** 10.1101/2024.03.15.24304164

**Authors:** Anh N. Do, Muhammad Ali, Jigyasha Timsina, Lihua Wang, Daniel Western, Menghan Liu, Jessie Sanford, Matitee Rosende-Roca, Merce Boada, Raquel Puerta, Ted Wilson, Agustin Ruiz, Pau Pastor, the Alzheimer’s Disease Neuroimaging Initiative (ADNI), Tony Wyss-Coray, Carlos Cruchaga, Yun Ju Sung

**Author notes:** Data used in preparation of this article were obtained from the Alzheimer’s Disease Neuroimaging Initiative (ADNI) database (adni.loni.usc.edu). As such, the investigators within the ADNI contributed to the design and implementation of ADNI and/or provided data but did not participate in analysis or writing of this report. A complete listing of ADNI investigators can be found at: http://adni.loni.usc.edu/wp-content/uploads/how_to_apply/ADNI_Acknowledgement_List.pdf.

## Abstract

In Alzheimer’s disease (AD), the most common cause of dementia, females have higher prevalence and faster progression, but sex-specific molecular findings in AD are limited. Here, we comprehensively examined and validated 7,006 aptamers targeting 6,162 proteins in cerebral spinal fluid (CSF) from 2,077 amyloid/tau positive cases and controls to identify sex-specific proteomic signatures of AD. In discovery (N=1,766), we identified 330 male-specific and 121 female-specific proteomic alternations in CSF (FDR <0.05). These sex-specific proteins strongly predicted amyloid/tau positivity (AUC=0.98 in males; 0.99 in females), significantly higher than those with age, sex, and APOE-ε4 (AUC=0.85). The identified sex-specific proteins were well validated (r≥0.5) in the Stanford study (N=108) and Emory study (N=148).

Biological follow-up of these proteins led to sex differences in cell-type specificity, pathways, interaction networks, and drug targets. Male-specific proteins, enriched in astrocytes and oligodendrocytes, were involved in postsynaptic and axon-genesis. The male network exhibited direct connections among 152 proteins and highlighted PTEN, NOTCH1, FYN, and MAPK8 as hubs. Drug target suggested melatonin (used for sleep-wake cycle regulation), nabumetone (used for pain), daunorubicin, and verteporfin for treating AD males. In contrast, female-specific proteins, enriched in neurons, were involved in phosphoserine residue binding including cytokine activities. The female network exhibits strong connections among 51 proteins and highlighted JUN and 14-3-3 proteins (YWHAG and YWHAZ) as hubs. Drug target suggested biperiden (for muscle control of Parkinson’s disease), nimodipine (for cerebral vasospasm), quinostatin and ethaverine for treating AD females. Together, our findings provide mechanistic understanding of sex differences for AD risk and insights into clinically translatable interventions.

## Introduction

Alzheimer’s disease (AD) is the most common cause of dementia, affecting over 6 million Americans and expected to rise over double (∼13 million) in 2050.^1^ AD has been a major topic of neurodegenerative research due to its high prevalence, increasing incidence, and lack of effective prevention.^2^ AD can be recognized by a plethora of symptoms including language difficulties and short-term memory deficits, and progressive loss of long-term memory and body functions.^2, 3^ Cellular hallmark of AD include cerebral plaques and synaptic disconnections. The cerebral plagues are composed of amyloid beta aggregations, which is derived from amyloid precursor protein.^2, 4^ Additionally, neurofibrillary tangles containing hyperphosphorylated tau are also observed in the brains of individuals with AD. The accumulation of amyloid beta is believe to initiate a cascade of events, leading to the abnormal phosphorylation of tau protein. This sequence of events, also known as the amyloid cascade hypothesis, is the current understanding for the clinical development and progression of AD. A latest anti-amyloid beta treatment appears to slow down cognitive decline, bringing new hopes for AD patients and their families, although long-term efficacy and safety remains to be seen.^5^

There are notable sex differences in the prevalence and progression of AD. Females have a higher risk of developing AD compared to males. Among 6.5 million seniors diagnosed with AD in US, around 4 million are females.^1^ Additionally, females face approximately double the risk of developing AD after the age of 45 compared to males in US.^6^ Worldwide AD prevalence and AD-related death in females are also higher compared to males (1.18 and 1.14 times, respectively, after adjusting for age).^7^ After diagnosis, females exhibit faster AD progression with more rapid cognitive and functional decline than males. In our 5-year longitudinal study of 1,499 individuals in the ADNI and Knight-ADRC cohorts, AD progression (i.e., clinical dementia rating) in females was 21% faster than males^8^. One common explanation is the higher life expectancy of females. However, this alone cannot account for the prevalence and risk difference fully.^9^ The knowledge about mechanism of sex-specific difference in AD are still limited.

Numerous genomic and transcriptomic studies have been conducted to elucidate the sex difference in AD at molecular levels.^9^ Multiple genome-wide association studies (GWAS) identified sex-specific variants associated with AD, amyloid pathology, tau pathology and soluble TREM2 in brain, cerebral spinal fluid (CSF), and plasma samples. Specifically, different genetic variants in serpin family members (*SERPINB1, SERPINB6,* and *SERPINB9*), which are involved in inflammatory response, were found to have strong associations with amyloidosis in females but not in males in a study of 3,036 brain and CSF samples.^10^ Other female-specific genetic associations in AD-related outcomes identified neurotrophin signaling pathway,^11, 12^ which regulates development, maintenance and function of nervous system, and genetic variants of *BDNF* which encodes a protein of the nerve growth factor family.^13^ In another study, genetic variants located within chromatin loops that interact with promoters of genes involved in RNA processing including GATA3 were reported having higher AD resilience scores in females.^14^ In males, strong AD signals were found for genetic variants of *GRN*, which encodes a protein acting as a key regulator of lysosomal function and as a growth factor involved in inflammation, *TREM2* regulating phagocytosis and cytokine productions, and *IGF2* involved in development and growth.^15-17^ In addition, several transcriptomic studies highlighted the critical roles of sex hormone genes including estrogen-related receptor beta in brain-sampled AD, sex steroids in hippocampal neuronal degeneration with androgen and estrogen receptors in homeostatic processes of AD neurons.^18 19^ Besides sex hormones, female-specific signatures seem to be enriched in immune response, inflammatory signaling pathways, and neuro-inflammation in AD in brain and blood.^9, 20-24^ Male-specific signatures were identified for expression of long noncoding RNAs, or nicotine addiction, adipo-cytokine signaling, and alcoholism as enriched pathways in association with AD in brain.^23, 25^

In contrast to the findings from genomic and transcriptomic studies, the impact of sex on proteomic signatures of AD are vastly understudied. Some small scale protein studies in blood reported female-specific results, including tropomyosin-1 protein that is significantly higher in AD females compared to AD or mild cognitive impairment (MCI) males and alpha-2 macroglobin that shows strong correlation with AD progression in females.^26 27^ In another study of 61 frozen basal forebrain samples, cortical histone deacetylase (HDAC2) protein levels in cholinergic neurons were significantly lower in females compared to males across disease status.^28^ A recent integrated proteomic and metabolomics study of 43 hippocampal samples also highlighted profound differences in insulin response and serine metabolism between control and AD males and females.^29^ A large-scale proteomic study of 1,277 samples from six brain regions found 1,239 proteins differed by sex in the dorsolateral prefrontal cortex, but biological pathways of the identified proteins were not reported.^30^ A limited number of these proteomic studies support a need for a large-scale and comprehensive proteomic study examining sex-specific differences in AD.

We hypothesized that males and females have different proteomic alterations by amyloid/tau positivity, which may in turn help explain their difference in AD prevalence. To test this hypothesis, we generated 7,000 proteomics data in CSF from 1,766 participants across four large and well-characterized cohorts. We pursued an unbiased and hypothesis-free approach to identify sex-specific proteomic signatures of AD pathology. We considered amyloid/tau positivity instead of clinical AD status for early diagnosis and potential intervention since changes in CSF amyloid/tau level changes occur a decade before clinical symptoms. In the downstream analyses, we validated our findings in external cohorts, identified the biological pathways, and built the interaction networks among proteins to elucidate their potential mechanism of actions for AD.

## Results

### Study overview and sample characteristics

To identify sex-specific effect of amyloid/tau positivity on CSF proteomics, we performed two-stage analysis: (1) sex-stratified analysis (males and females, separately) in the discovery stage with 1,766 individuals across four studies: the Knight Alzheimer Disease Research Center (Knight-ADRC), Alzheimer’s Disease Neuroimaging Initiative (ADNI), Bracelona-1, and Fundació Ace (FACE) studies and meta-analysis; (2) sex-stratified analysis in the validation stage with 256 individuals from the Stanford ADRC study and Emory Diversity study (Fig. 1a). CSF samples were collected from all individuals following a standard protocol. Amyloid/tau positivity was determined using amyloid beta 42 (Aβ42) and phosphorylated tau at threonine 181 (pTau) levels in CSF, based on the AT(N) classification.^31^ Briefly, quantitative Aβ42 and pTau measures were dichotomized into high and low levels based on a Gaussian mixture model, which was first described by our group.^10, 32^ Individuals with low CSF Aβ42 (A+) and high pTau levels (T+) were defined as amyloid/tau positive (A+T+) cases. Individuals with high Aβ42 (A-) and low pTau (T-) levels were defined as controls (A-T-). We obtained proteomic measurements using the SOMAscan platform for all four discovery cohorts and performed a standard data processing, normalization, and quality control, which resulted in 7,006 aptamers targeting 6,162 proteins available for analysis. In the validation cohorts, Stanford ADRC study used SOMAscan platform to quantify 4,735 analytes of 4,490 proteins, and Emory Diversity study used tandem mass tag mass spectrometry to measure 1,840 proteins. The validated male and female-specific proteins were separately followed by predictive models, biological pathways, interaction networks and drug targets (Fig. 1a).

**Figure 1:**
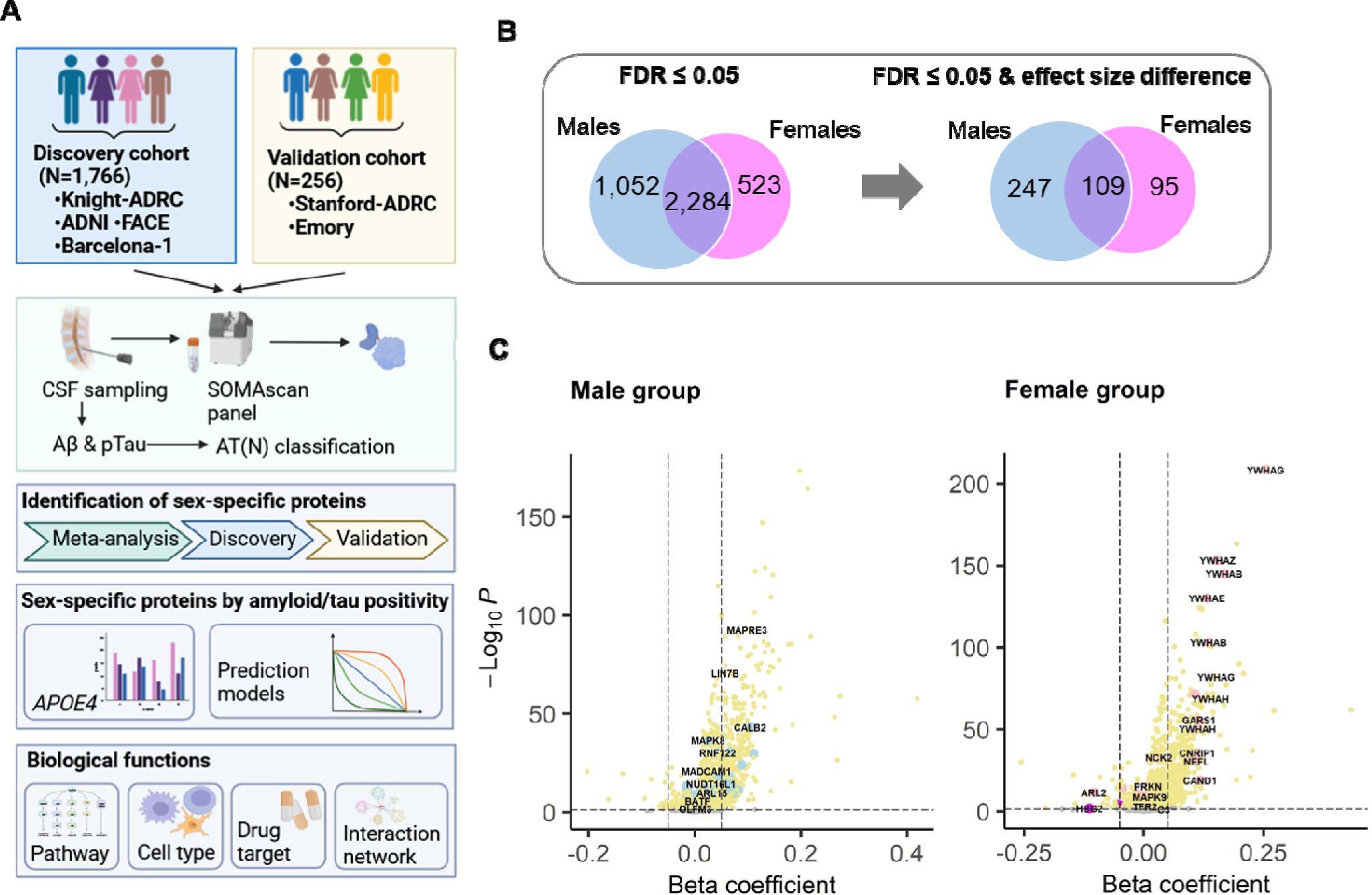
Study design and identification of sex-specific proteomic alteration by amyloid/tau positivity in discovery. (A) Study design. CSF samples of all individuals in the discovery and validation cohorts were quantified for protein levels. Amyloid/tau positivity was determined using AT(N) framework with CSF Aβ42 and pTau levels. First, we identified sex-specific proteomic alternation by amyloid/tau positivity in CSF. Second, we further examined the APOE ε4 modification effect and their predictive performance. Third, to uncover their biological functions, we performed pathway analysis, cell type enrichment, drug targets, and interaction network. (B) Sex-specific proteins identified in discovery follow the two-step strategy. The left panel showed number of proteins passed FDR≤0.05 in association with amyloid/tau positivity. Blue circles showed proteins significant in males. Pink circles showed proteins significant in females. The overlap showed proteins significance in both. The second panel showed number of proteins significantly associated with amyloid/tau positivity and having different effect size between males vs. females (P≤0.05). The same color coding was applied (C) Volcano plot of sex-specific proteins altered by amyloid/tau positivity in males and females identified in discovery. The left panel showed male-specific proteins identified in males. Blue dots show male-specific proteins. Yellow dots show proteins significance in males but having similar effect size between males and females. The right panel showed female-specific proteins identified in females. Pink dots show female-specific proteins. Yellow dots show proteins significance in females but having similar effect sizes between males and females. X-axis shows protein alterations by amyloid/tau positivity. Y-axis shows –log10 p-values of the associations.

The discovery cohort included 851 amyloid/tau positive cases and 915 controls, and the validation cohort included 84 cases and 178 controls (Table 1). Consistent to previous knowledge about risk factors of AD, cases were older and had more females and more *APOE* ε4 carriers, compared to controls. Individuals in validation were younger and had more proportion of females compared to those in discovery (Table 1).

**Table 1.**
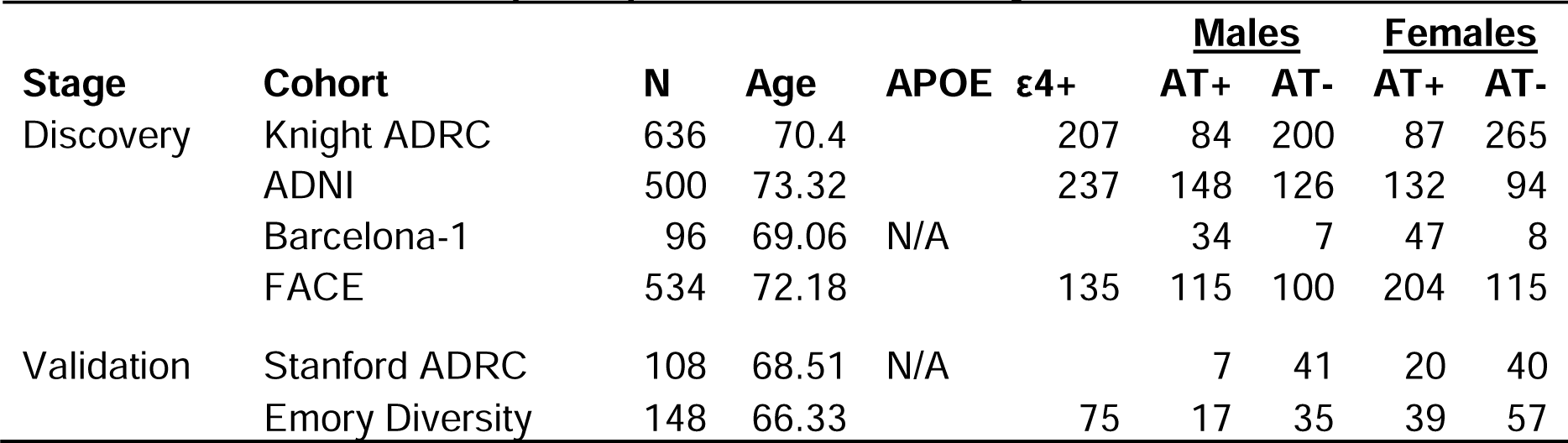
Characteristics of participants in the discovery and validation cohorts.

### Discovery of sex-different protein alteration by amyloid/tau positivity

To investigate the sex difference in the association of amyloid/tau positivity with protein levels, regression was performed. Log_10_ protein levels were used as a dependent variable and amyloid/tau positivity as an independent variable, while adjusting for age, cohort, and sample plate as covariates. The analysis was performed in males and females separately (sex-stratified data) and then compared the significance and effects of amyloid/tau positivity between two groups following three steps. First, we selected proteins that were significant in either sex (at FDR ≤ 0.05) and characterized them into three groups including significance only in males, only in females, and in both sexes. Second, we further selected proteins that had different their effect size of association with amyloid/tau positivity between males and females (P < 0.05). Third, we classified these proteins as male-specific if they were significant in males or if they had significantly stronger effects in males when significant in both sexes. Similarly, proteins were classified as female-specific if they were significant in females only or if they had significantly stronger effects in females when significant in both sexes.

In discovery, we identified 1,052 proteins that showed significant association with amyloid/tau positivity only in males, 523 proteins significant only in females, and 2,284 significant in both (FDR ≤ 0.05) (Fig. 1b and Fig. 1c). Among them, 247 were significant only in males and had significantly stronger effect size in males, referred as male-specific (dark blue in Fig. 2a). Of 95 proteins were significant only in females and had significantly stronger effect size in females, referred as female-specific (dark pink in Fig. 2a). Among proteins significant in both, 83 had stronger effect sizes in males, referred as male-specific (light blue in Fig. 2a) and 26 with stronger effect sizes in females referred as female-specific (light pink in Fig. 2a). In total, 330 proteins were male-specific and 120 were female-specific (Fig. 2a and Supplemental Table 1). The top ten sex-specific proteins ranked by P-value for their sex difference were shown in Fig. 2b. We found that female-specific proteins had stronger effect size compared to male-specific proteins (Fig. 2a and 2b). About 60% of sex-specific proteins showed increases in their abundances with amyloid/tau positivity.

**Figure 2:**
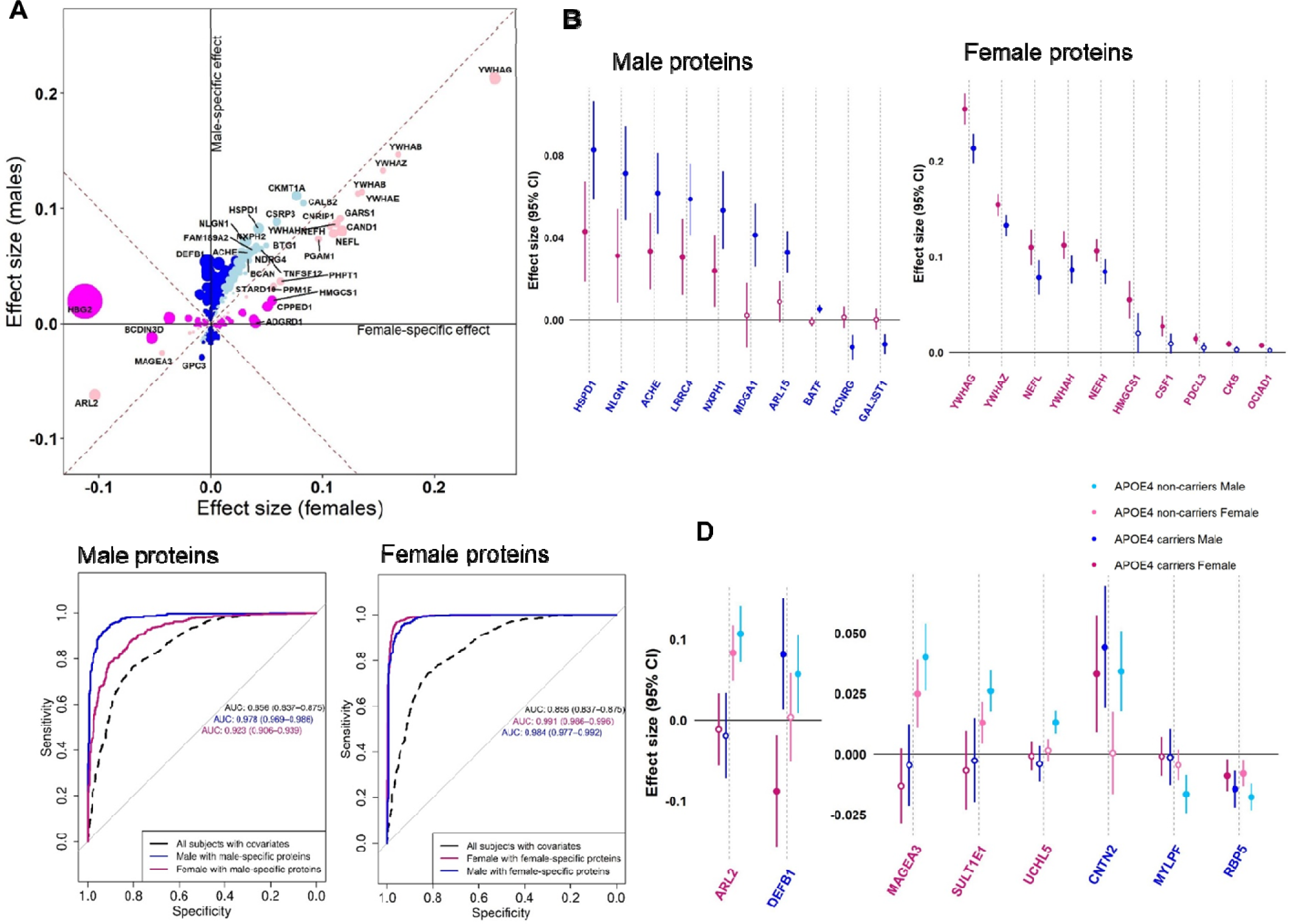
Sex-specific signature of proteins identified in discovery. (A) Sex-stratified 451 protein alterations by amyloid/tau positivity in discovery. There were 330 male-specific proteins (in blue dots) and 121 female-specific proteins (in pink dots). The dark blue dots show proteins significant in males only and having effect size stronger in males. The light blue dots show proteins significant in both and having effect size stronger in males. The dark pink dots show proteins significance in females only and having effect size stronger in females. The light pink dots show proteins significance in both and having effect size stronger in females. The size of dots shows magnitude of effect size different between sexes. X-axis shows protein alterations by amyloid/tau positivity in females, and y-axis shows those in males. (B) Forest plot of male and female protein alterations by amyloid/tau positivity in discovery. Y-axis shows beta coefficient with confidence interval of protein changes by amyloid/tau positivity in males and females, separately. Effects in males are in blue lines, and those in females are in pink lines. Male proteins are labeled in blue, and female proteins are in pink. (C) Predictive AUC curve of sex-specific proteins on amyloid/tau positivity. The left panel showed prediction performance of male-specific proteins. Black dashed curve is with basic models including age, sex and *APOE* genotype in all individuals. Blue curve is with 330 male-specific proteins in males. Pink curve is with 330 male-specific proteins in females. The right panel showed prediction performance of female-specific proteins. Black dashed curve is with basic models including age, sex and *APOE* genotype in all individuals. Pink curve is with 121 female-specific proteins in females. Blue curve is with 121 female-specific proteins in males. Y-axis shows sensitivity, and x-axis shows specificity. (D) Forest plot of protein changes by amyloid/tau positivity stratified by sex and *APOE* ε4. Y-axis shows beta coefficient with confidence interval of protein changes by amyloid/tau positivity in females and males, separately. Effects in male *APOE* ε4 carriers are in dark blue lines, and those in male *APOE* ε4 non-carriers are in light blue lines. Effects in female *APOE* ε4 carriers are in dark pink lines, and those in female *APOE* ε4 non-carriers are in light pink lines. Male proteins are labeled in blue, and female proteins are in pink.

### Validation of sex-different alteration by amyloid/tau positivity

For the proteins showing sex-specific effects in discovery, we sought to validate in two independent cohorts, Stanford ADRC study (N=108) and Emory Diversity study (N=148). Correlations of effect size and concordance of direction of effects between discovery and validation were investigated. The high correlation (r=0.74 in males and r=0.82 in females) of effect sizes between discovery and Stanford ADRC study (P<2.2×10^-16^) among 4,647 overlap proteins were observed (Supplemental Fig. 1). We found moderate correlation (r=0.47 for males and r=0.59 for females) of effect sizes between discovery and Emory Diversity study (P<2.2×10^- 16^) among 1,267 overlap proteins (Supplemental Fig. 1), which is expected as orthogonal proteomic platforms were used (SOMAscan in discovery vs. mass spectrometry in Emory study). Additionally, the kappa statistics showed a strong agreement of direction of effects between proteins in discovery and Stanford ADRC study (kappa=0.87 in males; kappa=0.85 in females; Supplemental Table 2). The agreement of direction of effects between discovery and Emory Diversity study was decent (kappa=0.67 in males; kappa=0.61 in females; Supplemental Table 2).

### Predictive performance of sex-specific proteins

To evaluate the predictive performance of the 451 sex-specific proteins on the amyloid/tau positivity, we created prediction models for amyloid/tau positivity in sex-stratified groups by using discovery and then validated in the validation. Because age, sex, and *APOE* genotype are strong risk factors of AD^2^, we included them as covariates in a basic model for reference. Sex-specific prediction models were built using sex-specific proteins (no other covariates included) for the corresponding sex-stratified groups. Stepwise regression was applied to select a subset of sex-specific proteins in the model. Among 330 male-specific proteins, 64 were included in the prediction model after stepwise regression (Supplemental Table 3). The model based on the 64 male-specific proteins provided significantly higher predictive performance for amyloid/tau positivity in males when compared to the basic model, with area under the curve (AUC) = 97.8% vs. 85.6% (P = 2.20×10^-16^, Fig. 2c). Among 121 female-specific proteins, 49 were included in the prediction model for amyloid/tau positivity after stepwise regression (Supplemental Table 3). The model based on the 49 female-specific proteins provided significant higher predictive performance for amyloid/tau positivity in females when compared to the basic model with age, sex, and *APOE* genotype (AUC = 99.1% vs. 85.6%; P = 2.20×10^-16^). The results were validated in the Stanford ADRC (AUC=95.6% in males with 37 proteins; 97.9% in females with 29 proteins) and Emory Diversity studies (AUC=74.5% in males with 24 proteins; 98.9% in females with 12 proteins; Supplemental Fig. 2).

As a sensitivity analysis, these sex-specific proteins remained after stepwise regression were used to build prediction models in the other sex group (e.g., applying male-specific prediction to female group). The male-specific model predicted significantly lower in females (AUC=92.3%, Fig. 2c) indicating that they were optimized for the prediction in males. On the other hand, the female-specific model predicted well for males (AUC=98.4%, Fig. 2c).

### Pronounced sex-specific alterations in *APOE* carriers

Several publications suggested that *APOE* ε4 genotype may modify sex-specific effect on the prevalence and progression of AD^33-36^. To explore the potential relationships between sex and *APOE* ε4 status, we performed sex-specific and *APOE* ε4 interaction association analyses with amyloid/tau positivity in discovery for the 451 sex-specific proteins. Among sex-specific proteins, four were differently associated with amyloid/tau positivity across sex and *APOE* ε4 genotype groups (FDR≤0.05) and 66 proteins had marginal effects (P≤0.05) (Supplemental Table 4). Fig. 2d exhibited eight female/male-specific proteins (ranked by p-value) associated with amyloid/tau positivity across different sex and APOE ε4 groups. Within *APOE* ε4 carriers, DEFB1, a male-specific protein had a positive association with amyloid/tau positivity among males but a negative association among females which was not observed in *APOE* ε4 non-carriers (Fig. 2d). Four other female-specific proteins (ARL2, MAGEA3, SULT1E1, and UCHL5) and three other male-specific proteins (CNTN2, MYLPF, and RBP5) had a strong effect difference between sexes among *APOE* ε4 non-carriers but not among *APOE* ε4 carriers (Fig. 2d).

### Biological pathways

To uncover biological processes enriched in the identified proteins, we performed gene ontology (GO) and pathway analysis using Enrichr^37^ for male-specific and female-specific proteins separately. The 330 male-specific proteins were enriched for synaptic structures (fold enrichment >20; FDR≤0.01) and axon activities (fold enrichment=8.2; FDR=2.0×10-4) (Fig. 3a and Supplemental Table 5). We obtained clustering of the identified GO biological processes based on their similarity calculations to distill them into common higher-level representative terms using REVIGO^38^. Interestingly, all of these GO terms were clustered in higher-level terms of nervous system development and axonogenesis, revealing distinctive sex-specific biological processes (Fig. 3b). Notably, male-specific proteins were highly enriched in genome-wide association phenotypes including cognitive ability, rate of cognitive decline in AD, schizophrenia (fold enrichment>2; FDR≤0.05) (Fig. 3c and Supplemental Table 6). The 121 female-specific proteins were enriched for different receptor bindings including nuclear glucocorticoids, phosphoserine, and cytokine (fold enrichment >3; FDR≤0.05) (Fig. 3d and Supplemental Table 7). We found phosphoserine residue binding and DNA-binding transcription activator activity as higher-level GO terms for the female-specific proteins (Fig. 3f). Pathway analysis revealed that female-specific proteins were involved in inflammatory processes including oxidative stress response and T-cell activation as the top enrichments (Fig. 3g and Supplemental Table 8). These results taken together demonstrated that male and female-specific proteins were involved in different biological processes suggesting potential different pathophysiology of AD in sex groups.

**Figure 3:**
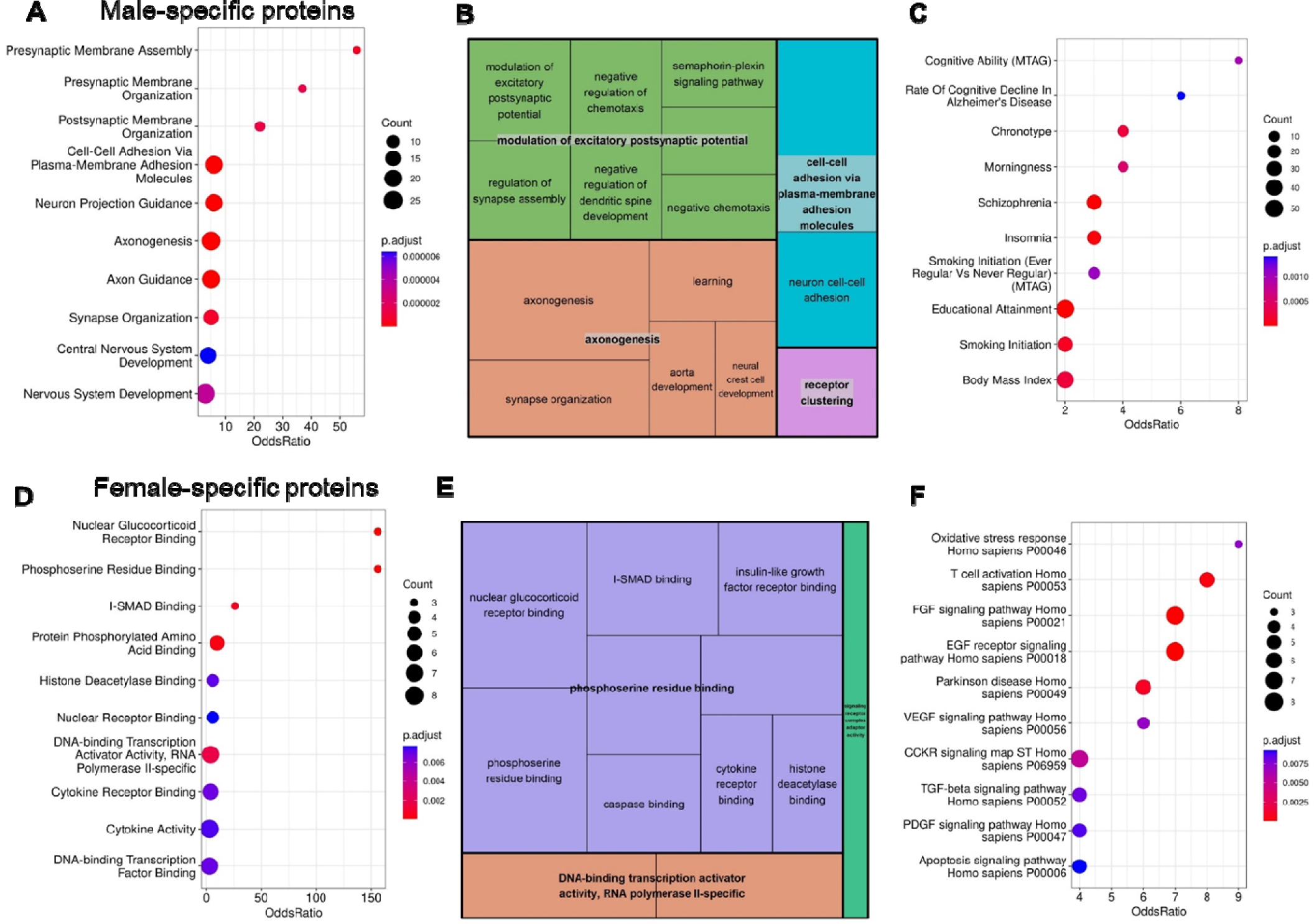
Gene ontology (GO) biologic processes of sex-specific proteins. (A) Enrichment analysis of 330 male-specific proteins in discovery revealed presynaptic membrane assembly as the most enriched for these proteins. Top gene ontology (GO) terms (FDR≤0.05) are ranked by odd ratios. Row name shows GO terms. (B) Graphical REVIGO schematic of GO terms (FDR≤0.05) for 330 male-specific proteins reveals modulation of excitatory postsynaptic potential and axonogenesis as the most enriched biological processes. Box size inversely corresponds to FDR for association. Bolded terms indicate higher order GO processes with the consistent color across the GO terms per each higher order term. (C) Enrichment analysis of 330 male-specific proteins in discovery revealed cognitive ability as the most enriched for these proteins. Top gene ontology (GO) terms (FDR≤0.05) are ranked by odd ratios. Row name shows GWAS phenotypes from GWAS catalog. (D) Enrichment analysis of 121 female-specific proteins in discovery nuclear glucocorticoid receptor binding as the most enriched for these proteins. Top gene ontology (GO) terms (FDR≤0.05) are ranked by odd ratios. Row name shows GO terms. (E) Graphical REVIGO schematic of GO terms (FDR≤0.05) for 121 female-specific proteins reveals modulation of phosphoserine residue binding as the most enriched biological processes. Box size inversely corresponds to FDR for association. Bolded terms indicate higher order GO processes with the consistent color across the GO terms per each higher order term. (F) Enrichment analysis of 121 female-specific proteins in discovery revealed oxidative stress response and T-cell activation as the most enriched for these proteins. Top gene ontology (GO) terms (FDR≤0.05) are ranked by odd ratios. Row name shows human biological pathways.

### Cell type specificity

To examine the cellular context of the identified proteins, we used cell type expression data from human brain cells.^39^ For the 330 male-specific proteins, cell type estimation was available for 300 proteins, in which 103 had a specific cell type with abundance over 50%. They were involved in cells of central nervous system: astrocytes (24 proteins), microglia (22 proteins), neurons (39 proteins), and oligodendrocytes (10 proteins) (Supplementary Fig. 3). The 121 female-specific proteins were involved in cells of central nervous system including astrocytes (3 proteins), microglia (7 proteins), neurons (14 proteins), and oligodendrocytes (1 proteins) (Supplementary Fig. 4). There is a significant difference between cell types in male-specific proteins compared to those in female-specific proteins (P of Chi-square test=0.02) especially for astrocytes and oligodendrocytes (Fig. 4a). Astrocytes and oligodendrocytes were highly enriched for male-specific proteins (over enriched; fold enrichment=1.62 and 1.46, respectively) but not for females-specific proteins (under enriched; fold enrichment=0.55 and 0.39, respectively).

**Figure 4:**
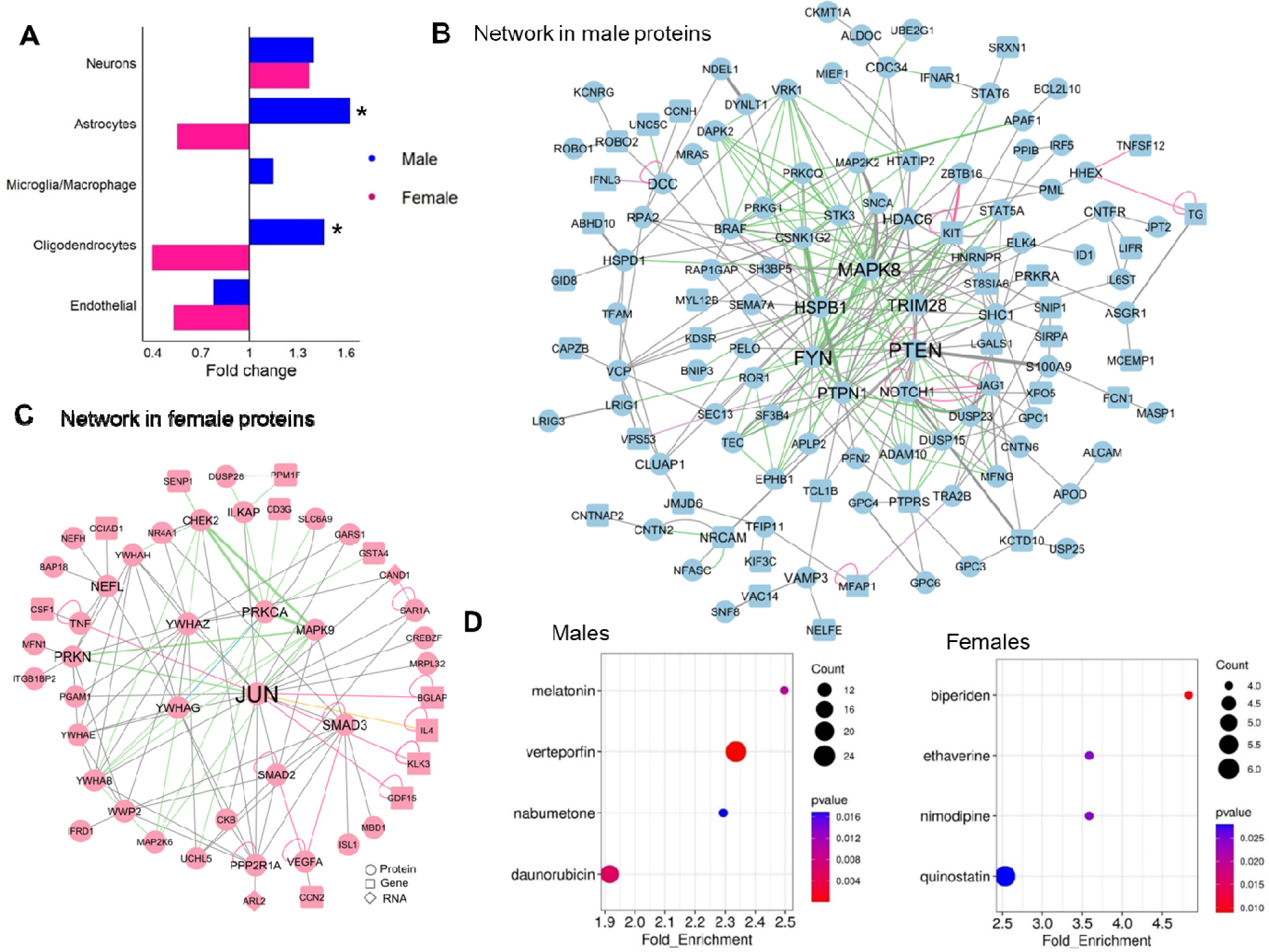
Functional characterization of the sex-specific proteins. (A) Cell types enriched for 451 sex-specific proteins. Blue bars show male-specific proteins. Pink bars show female-specific proteins. The black vertical line shows fold enrichment at 1 to separate over or under enrichment. X-axis shows fold enrichments. Row name shows cell types. The star indicates the significant difference of cell type enrichment between sexes (P<0.05). (B &C) Molecular interaction networks of male-specific proteins and female-specific proteins from ConsensusPathDB, respectively. Blue dots show male-specific proteins. Pink dots show female-specific proteins. Circle shapes indicate proteins. Rectangle shapes indicate genes. Diamond shapes indicate RNA. The size of dots shows the number of other dots directly connecting to the dots. The thickness of edges shows the number of prior publications supporting the interactions. Green edges indicate biochemical interactions. Pink edges indicate gene regulatory interaction. Grey edges indicate protein interactions. Hubs of the networks with more interactions to other genes/proteins were shown in the middle of the network. (D) Drug target prediction using the sex-stratified effects of the 451 sex-specific proteins with left panel for male-specific proteins in males and right panel for female-specific proteins. Top drug targets (FDR≤0.05) are ranked by odd ratios. Row name shows drug names.

### Interaction network of sex-different proteins

To identify potential interactions among the 451 identified proteins, we constructed interaction networks for male and female-specific proteins, separately, using ConsensusPathDB.^40^ The network for male-specific proteins showed the inter-connections among the 152 proteins (Fig. 4b). PTEN, FYN, and MAPK8 were hubs of the network which directly connected 31, 30, and 28 proteins/genes, respectively. Among three hubs of the network, PTEN levels were significantly associated with lower risk of AD observed only in males, whereas two other proteins (FYN and MAPK8) were associated with higher risk of AD in both males and females but with stronger effect in males. The male-specific network had majority of protein-protein interaction (181; 58.4%), followed by biochemical reaction (113; 36.5%), gene regulatory interaction (5.2%), and genetic interaction (1.6%). The network for female-specific proteins showed the inter-connections among the 51 proteins (Fig. 4c). In particular, as a hub of the network, a transcription factor JUN showed its connection with 30 different proteins/genes. JUN levels were significantly associated with higher risk of AD among females but not observed in males. Similar to male-specific network, majority (70; 53.9%) of them were protein-protein interaction, followed by biochemical reaction (28; 21.5%).

### Drug targets

To explore potential drug targets that have effects on AD through the 451 sex-specific proteins, we used Connectivity Map dataset containing of 7,056 transcriptomic data of 1,310 drug perturbations through Ensemble of Multiple Drug Repositioning Approaches (EMUDRA).^41^ Using male-specific protein alteration by the amyloid/tau positivity, we found melatonin, vertoporfin, nabumetone, and daunorubicin as drug targets for AD (fold enrichment ≥1.9 and P < 0.05; Supplemental Table 9; Fig. 4d). Verteporfin is a benzoporphyrin derivative and used as a photosensitizer in photodynamic therapy to eliminate abnormal blood vessels in wet form macular degeneration^42^. Verteporfin was predicted to regulate 24 different genes, some of which were involved in neural activities including BAMBI, GPC4, JAG1, LRP4, LRIG1 and ROBO1. Daunorubicin is an anthracycline antibiotic, mainly used in cancer treatment, particularly for different types of leukemia.^43^ However, the effects of these two drugs on AD have not been established. The use of melatonin and nabumetone in AD treatment has been initiated. Melatonin is an endogenous hormone secreted from pineal gland, primarily known for its role in regulating sleep-wake cycles, and used off-label for insomnia. Additionally, melatonin has been proposed as a potential treatment for AD-related dementia. However, more research is needed to confirm^44^. Nabumetone is a nonsteroidal anti-inflammatory drug, commonly used for mild to moderate pain and reduces symptoms of arthritis.^45^ While there is some suggestion that nabumetone may have benefits in reducing the risk of AD,^46^ further studies are needed to fortify the findings.

Similarly, using female-specific protein alteration by the amyloid/tau positivity obtained from female-only analysis, we found biperiden, ethaverine, nimodipine, and quinostatin as drug targets predicted to have effects on AD (Supplemental Table 9 and Fig. 4b). Biperiden is a muscarinic cholinergic receptor antagonist and commonly used to treat the symptoms of Parkinson’s disease including stiffness, tremors, and power muscle control.^47^ Ethaverine is an effective smooth muscle relaxant that acts by relaxing the smooth muscles in various areas of the body including coronary arteries, cerebral arteries, pulmonary arteries, and peripheral arteries.^48, 49^ However, their effects on AD have not been established. Nimodipine is a second-generation calcium channel blocker that has been approved by the FDA for treatment and prevention of cerebral vasospasm.^50, 51^ Regarding cognitive impairment and dementia, while nimodipine may be used off-label for these conditions in some countries, it does not have FDA approval for these indications.^50, 51^ Quinostatin is a chemical compound known to inhibit the lipid-kinase activity of the catalytic subunits of class I-a PI3Ks. Its potential as a treatment of cancer is currently being investigated.^52^

## Discussion

This is one of the first studies systematically investigating the influence of sex on protein alterations by amyloid/tau positivity in a large scale. By comprehensively examining over 7,000 proteins in CSF from 2,077 individuals, we pursued an unbiased and hypothesis-free approach to identify the proteins that were altered by amyloid/tau positivity in a sex specific manner. CSF is neurologically relevant tissue and effectively recapitulates neurological disorders compared to plasma. CSF samples from living participants have diagnostic values compared to brain samples from deceased. We examined amyloid/tau positivity based on A-T-N framework with CSF Aβ42 and CSF p-tau levels that are well-known to change decades before clinical symptoms appear. By employing statistical models in a sex-stratified manner, we discovered significant differences between males and females in their protein alterations by amyloid/tau positivity. We identified 330 male-specific proteins showing more pronounced changes in males and 121 female-specific proteins. The identified proteins were validated in two external studies with moderate to high correlations. Pathway analysis identified male-specific proteins enriched for nervous system development and axongenesis and female-specific proteins enriched for inflammatory processes, revealing distinctive sex-specific biological processes.

*APOE* is the strongest genetic risk factor for AD and may modify sex-specific alterations by amyloid/tau positivity. Among identified proteins, sex-specific alterations could be pronounced based on *APOE* ε4 information, suggesting that these sex differences were intertwined with *APOE* genotype. DEFB1, a male-specific protein showed strong alterations among *APOE* ε4 carriers in which protein levels increased with amyloid/tau positivity in males but decreased in females. This was observed only in *APOE* ε4 carriers. Several male-specific proteins (top three proteins, CNTN2, MYLPF, and RBP5, ranked by P-values in their sex differences) showed strong alterations among *APOE* ε4 non-carriers compared to carriers. Top four female-specific proteins (ARL2, MAGEA3, UCHL5, and SULT1E1) also exhibited the same pattern. Our results highlight the importance of examining modification effects of sex and *APOE* genotype, which help identifying proteins underpinning of AD that may be beneficial to specific patient groups.

The disease prediction models suggest that the 451 identified proteins provide high predictive performance for amyloid/tau positivity. Using male-specific and female-specific proteins alone could strongly predict AD with high predictive performance for each sex group without age, sex, and *APOE* genotype. Interestingly, the male-specific proteins also showed good performance with much higher predictive performance in males compared to females. Compared to previous studies, the proteins identified in this study appear to have higher prediction values. In a whole blood transcriptomic study, classification of AD built from support vector machine revealed AUC of 0.8 for males and 0.91 for females even with demographic information (age, sex, education, and *APOE* ε4) and gene expression levels.^20^ The identified proteins predicted AD better than well-established blood-based biomarkers including Aβ42 and pTau with the highest AUC=0.91.^53-56^ Taken together, the prediction models using CSF proteomics have significant clinical values as they are based on CSF from living individuals and seem to outperform blood-based biomarkers. Therefore, our sex-specific proteins identified here may be valuable as potential AD biomarkers.

The pathway analysis revealed the distinctive sex-specific biological processes. Male-specific proteins were involved in nervous system development with presynaptic membrane assembly and organization as top processes. Alterations in presynaptic membrane functions or structures including calcium signaling,^57^ cholesterol homeostasis,^58^ presynaptic proteins in the outer molecular layer of the dentate gyrus,^59^ presynaptic membrane proteins are reported to be associated with AD. A previous sex-specific transcriptomic difference study in brain found genes associated with male AD were enriched in pre-synapse and ATP metabolic process^9^. In contrasts, female-specific proteins were involved in inflammatory response with nuclear glucocorticoid receptor binding, oxidative stress response, and T-cell activation as the top processes. The roles of glucocorticoid receptor and T-cell in AD pathology has been studied. Prior studies showed that elevated glucocorticoid hormone levels enhanced Aβ levels which causes the impaired cognition in murine.^60, 61^ An experiment that blocked glucocorticoid receptor for only three days found a tremendous decrease of Aβ40 and Aβ42 in hippocampus and cognitive deficits recovery in mice.^61^ A recent study found a much increased number of T cells in tau pathology mice and in the human Alzheimer’s disease brain.^62^ Our pathway findings in female group overlapped with a prior meta-analysis study of sex-specific DNA methylation difference using over 1,000 brain samples which identified female-specific CpG sites involving in TYROBP causal network with TCR signaling in naïve CD4L+LT cells.^63^ Another sex-specific transcriptomic difference study in brain found genes associated with female AD involved in response to growth factor and response to wounding which glucocorticoid receptors play a critical role.^64^ Our validated findings here help uncover sex differences in these pathways related to AD pathogenesis.

Our interaction network suggests that the central hub of the network for males, PTEN, may be served as a target for AD therapeutics. PTEN is a functional kinase antagonizing the PI3K-AKT/PKB signaling pathway which plays an important role in mitochondrial energetic metabolism by promoting cyclooxygenase activity and ATP.^65^ Mutations in PTEN are associated with multiple neurological disorders including autism and seizures.^66^ In AD, PTEN was shown to act as a downstream of Aβ in a common pathway to depress synaptic transmission.^65^ PTEN was also found to mediate the synaptic plasticity that contributes to cognitive processes and normal brain function.^67^ Notably, blocking the recruitment of PTEN to synapses was shown to possibly reduce the onset and delay progression of neuronal and cognitive deficits in AD patients.^65^ We found JUN as a central hub of the network for females. JUN is a transcription factor that recognizes and binds to the AP-1, another transcription factor to regulate gene expression of multiple stimuli including cytokines, growth factor and stress.^68^ JUN plays a critical role in activation-induced cell death of T-cell by binding to the AP-1 promoter site.^68^ A recent study found that upregulation of JUN caused to increase the accessibility of genomic regions containing transposable elements which initiated cell death and activated immune response in neurons.^69^ Interestingly, inhibiting JUN rescued neuronal death and reversed impaired neurogenesis in the AD hippocampal progenitors.^69^ Our findings on human aligned with these results and confirmed JUN as a potential therapeutic strategy and biomarker for AD. These findings together with our results on human may pave a new avenue for AD prevention and treatment.

Our drug target prediction suggests potential drugs for AD in sex-specific way. In males, nabumetone may be a promising drug for AD males based on its anti-inflammatory effects through inhibiting the enzyme cyclooxygenase. A longitudinal study of over 5,000 elderly in Spain found that using nabumetone or similar drugs in its class was associated with 71% lower risk of AD mortality compared to not using.^45^ A recent meta-analysis study using Mendelian randomization to control for unknown confoundings confirmed these benefits on lowering AD risk, although the study also suggested the effect may be influenced by coronary heart disease, blood pressure, and blood lipids.^70^ Besides nabumetone, melatonin may be a potential drug of AD in males which was reported to enhance hippocampal neurogenesis and improve memory function.^71^ A meta-analysis of twenty-one clinical trials concluded that melatonin was beneficial in treatment of mild stage of AD with the significant improve of mini-mental state examination after 12 weeks of treatment.^44^ The mechanism of melatonin underlying these effects through balancing circadian rhythmicity, regulation of the immune system, and antioxidant effect.^71^ However, melatonin has not been approved by FDA for any usage on AD. Nimodipine suggested for females in our study has been reported to improve cognitive functions in different studies. A meta-analysis of fourteen clinical trials evaluating clinical efficacy of nimodipine on dementia concluded that patients on nimodipine had significant improved cognitive function, clinical global impression, and had a low rate of adverse effects compared to placebo group.^50^ The mechanism action of nimodipine on AD is not fully established, although a prior study found its action through its anti-inflammatory effects as nimodipine inhibited Aβ-stimulated IL-1β synthesis and release from microglia cell lines.^72^ In contrast to the positive findings, we also found contradictory results on biperiden which were opposite to previous publications. Biperiden was reported to impair memory performance in three clinical trials including decreasing the number of words recalls, delayed recall of the verbal learning task, reduced verbal memory, adaptive tracking, recognition memory in healthy volunteers at different ages.^47, 73, 74^ More studies are needed to further investigate the effect of drugs on sex-specific and AD-related outcomes to validate the results.

Our study has some limitations. First, while this study has the largest sample size to examine sex-specific CSF proteomics as of today, examining the differences between two groups (males vs. females) is known to be challenging, and therefore our study is relatively under powered to detect sex-specific signals. Second, medication use including hormone therapy may have influenced protein levels, so our findings may in part reflect some of medication use. This is a common limitation to many AD studies when medication information is not available in the database or it is not ethical to ask individuals especially seniors to stop medication use for study participation. Third, our study does not have information about cognitive status including subjective cognitive decline, MCI, and dementia however this study used amyloid/tau positivity as an outcome which were determined by CSF Aβ42 and pTau for early diagnosis and potential intervention as their changes occur a decade before clinical symptoms. Fourth, recent Lecanumab study identified improvement of the clinical dementia rating and several cognitive scores in sex-specific way. Linking our results in relationship to the finding from the study requires an additional follow-up

In summary, we identified and validated 330 male-specific proteins and 121 female-specific proteins in multiple cohorts. We examined these proteins showing sex different alterations by amyloid/tau positivity in biological processes and interaction network. The identified proteins may represent potential therapeutic targets for mitigating AD. Nimodipine and nabumetone, two FDA approved drugs as well as melatonin and their protein targets can be further studied to understand drug mechanisms in sex-specific manner. Together, our findings provide mechanistic understanding of sex differences for AD risk and insights for clinically translatable interventions.

## Methods

This study was approved by the Institutional Review Boards of the Washington University School of Medicine in St. Louis and the research was performed in accordance with the approved protocols.

### Study populations

This study examined 1,766 unique individuals with proteins measured by immunoassays and SOMAscan from four different cohorts: Alzheimer’s Disease Neuroimaging Initiative (ADNI), FACE, Charles F. and Joanne Knight Alzheimer Disease Research Center (Knight ADRC) and Barcelona-1. The cross-sectional study design was used in this project which included common samples between the platforms based on individual ID and CSF draw date match.

#### Knight ADRC

Charles F. and Joanne Knight Alzheimer Disease Research Center (Knight ADRC), housed at Washington University in St. Louis, is one of 30 ADRCs funded by NIH. The goal of this collaborative research effort is to advance AD research with the ultimate goal of treatment or prevention of AD. The individuals included in this study are from the Memory and Aging Project (MAP) supported by Knight ADRC. As part of the project, individuals undergo annual psychometric testing and interviews along with biennial or triennial PET, MRI and CSF collection. Further details on Knight ADRC and MAP can be found at https://knightadrc.wustl.edu/

#### ADNI

Data used in the analyses performed in this article were obtained from the Alzheimer’s Disease Neuroimaging Initiative (ADNI) database (adni.loni.usc.edu). ADNI was launched in 2003 as a public-private partnership, led by Principal Investigator Michael W. Weiner, MD. The primary goal of ADNI has been to test whether serial magnetic resonance imaging (MRI), positron emission tomography (PET), other biological markers, and clinical and neuropsychological assessment can be combined to measure the progression of MCI and early Alzheimer’s disease (AD). For up-to-date information, see www.adni-info.org.

#### Fundació ACE

The cerebrospinal fluid samples were obtained from Fundació ACE, a private non-profit group dedicated to the study of Alzheimer’s disease. Headquartered in Barcelona, Fundació ACE was founded in 1995 and has collected and analyzed almost 18,000 genetic samples, diagnosed over 8,000 patients, and participated in almost 150 clinical trials during its existence. For more details, visit www.fundacioace.com/en.

#### Barcelona-1

Barcelona -1 is a longitudinal observational study consisting of ∼300 individuals at baseline carried out in the Memory and Disorder unit at the University Hospital Mutua de Terrassa, Terrassa, Barcelona, Spain. Cases include individuals diagnosed with AD dementia, non-AD dementias, MCI, or individualive memory complaints (SMC). Clinical information was collected at baseline as well as longitudinally and lumbar puncture (LP) and amyloid PET were performed if individuals had diagnosis of MCI, early-onset dementia (<65 years), or dementia with atypical clinical features.

#### *Stanford ADRC* (Stanford Iqbal Farrukh and Asad Jamal ADRC)

Samples were acquired through the National Institute on Aging (NIA)-funded Stanford Alzheimer’s Disease Research Center (SADRC). The SADRC cohort is a longitudinal observational study of clinical dementia subjects and age-sex-matched non-demented participants. The study includes healthy control participants who were determined to be cognitively unimpaired through a clinical consensus conference involving board-certified neurologists and neuropsychologists. Cognitively impaired participants underwent Clinical Dementia Rating and standardized neurological and neuropsychological assessments to determine cognitive and diagnostic status, including procedures of the National Alzheimer’s Coordinating Center (https://naccdata.org/). All participants included in this study were deemed cognitively impaired during a clinical consensus conference that included neurologists and neuropsychologists. All participants were free from acute infectious diseases and in good physical condition. CSF samples was collected by lumbar puncture based on a standard protocol.

#### Emory Diversity

All CSF samples were collected as part of ongoing studies at Emory’s Goizueta Alzheimer’s Disease Research Center (ADRC). Clinical history, neurological examination, detailed cognitive testing, and diagnostic studies including magnetic resonance imaging as well as CSF AD biomarker testing were used to assess the clinical diagnosis of AD^75^. A consensus clinical diagnosis of controls was made without consideration of CSF biomarkers by a panel of experts at the Emory Goizueta ADRC^75^. The control participants were considered to have normal cognition and normal behavior after reviewing all testing including Montreal Cognitive Assessment (MoCA), Clinical Dementia Rating (CDR) score, and detailed neuropsychological testing. Control participants therefore may have MoCA scores that are lower than traditional cut points for impairment on this screening test^75^. CSF samples was collected by lumbar puncture based on the National Institute on Aging for Alzheimer’s Disease Centers guidelines (https://alz.washington.edu/BiospecimenTaskForce.html). Measurements of Aβ42, tTau, and pTau181 were performed on the Roche Diagnostics Elecsys platform based on the recommended protocols^75^. In total, the cohort had 105 healthy controls and 98 AD.

### Outcome definition based on AT(N) classification framework

Aβ42 and pTau biomarker levels from CSF samples were used to classify participants into cases and controls based on AT(N) classification framework which was applied separately for each cohort and separately for Aβ42 and pTau biomarkers.^31^ All samples in four cohorts were undergone the identical procedures as presented below.^32^ First, missing values were carefully handled. If a platform had many missing samples and other platforms were available, missing samples were rescued by using another platforms. The raw values of each biomarker were combined from two platforms based on the availability and high corrections among platforms. Samples in the Knight-ADRC cohort were went through this step due to data availability. Second, log10 transformation of AB42 and pTau biomarkers were conducted to make the biomarker distribution close to normal distribution. Then z-score was applied to standardize the distribution. Third, outliers were removed using ± 3×standard deviation cutoff. A total of 2 samples of Aβ42 (0.08%) and 7 samples of pTau (0.28%) were excluded. Fourth, standardization was applied to all samples in each cohort since mean of z-score changed as outliers were detected and removed. Finally, dichotomization using Mclust function “mclust” R package (version 5.4.6) via Gaussian mixture models fitted by expectation-maximization algorithm.^32^ After implementing all steps, quantitative biomarker levels of AB42 and pTau were classified into two groups corresponding to high and low groups. Samples with Aβ42 and pTau (A+T+) were defined as AD risk cases, while Aβ42 and pTau (A-T-) were defined as controls. A total of 851 amyloid/tau positive cases and 915 controls in discovery as well as 84 cases and 178 controls were defined in validation. For the sensitivity analysis, 525 amyloid positivity tau negativity (A+T-) were included as preclinical phase of AD in discovery. Only A-T+ was excluded as its unknown biological meanings. Notably, a small number of subjects were classified as A-T+ in both discovery and validation. Specifically,108 A-T+ (compared to 1,766 A+T+ and A-T-) were excluded from the discovery cohort with 67 A-T+ in Knight ADRC, 25 in ADNI, and 16 in Barcelona-1. A total of 12 A-T+ (compared to 311 A+T+ and A-T-) were excluded from the validation cohort with 6 A-T+ in Stanford ADRC and 6 in Emory Diversity. For the clinical diagnosis, case-control status was determined by the Clinical Dementia Rating (CDR) at the time of lumbar puncture which was used as clinical AD status in this project.

### Proteomics data collection, preparation, data processing and quality control (QC)

CSF samples of four cohorts in discovery including ADNI, FACE, Knight ADRC and Barcelona-1 were collected using lumbar puncture in the morning after an overnight fast. Samples were undergone the identical protocols for preparation and processing. All samples were processed and stored at -80 °C. All samples were then sent together to SomaLogic and were randomized across plates to avoid batch effects. Protein levels were measured using the SomaLogic aptamer-based SOMAscan platform based on a multiplexed, single-stranded DNA aptamer assay. The quantitative levels of 7,006 aptamers in 6,139 protein targets were reported as relative units of intensity (RFU or Relative Fluorescence Unit). Initial data normalization was performed by SomaLogic using hybridization controls for intra-plate and median signal to account for inter-plate variances.^76^ Normalization against an external reference to control for biological variances was also performed by SomaLogic.^77^ Individual level QC was then performed to exclude outliers. More details of data QC were published elsewhere.^77, 78^ A total of 1,766 samples with 7,006 aptamers were included in this project.

### Identification of AD sex-specific proteins

To identify sex-specific proteomic signatures of AD risk, CSF proteomic profiles from the discovery cohort were used to construct linear regression models using the *lm* function in the *stats* package in R. Protein levels were log10-transformed to make their distributions close to normal distribution. Age, sex, cohort, and sample plates were included as covariates. The analysis was performed in males and females separately (sex-stratified data). The results were then compared in term of the significance and effects of amyloid/tau positivity between two groups to identify proteins associated with AD risk and difference between females and males.

log_10_(protein level) ∼ AD risk + age + cohort + sample plates

P values of AD risk from the model in each sex group was reported. The results from studies in discovery were meta-analyzed using METASOFT 2.0.1^79^ separately for each sex group. Fixed effects model based on inverse-variance-weighted effect size and its corresponding p-values were used if FDR of P values of Q-test >0.05; otherwise, random effects model was included. Proteins were considered significant if FDR ≤ 0.05. Effect sizes of proteins in the association with amyloid/tau positivity were compared between male and female groups based on z-statistics.^80^

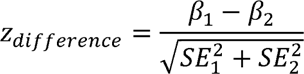

P-value ≤0.05 was considered as having effect size difference between sexes. Proteins passing both above defined significance thresholds (FDR ≤ 0.05 and P < 0.05 for effect size difference) were considered as “sex-specific” proteins. The proteins were classified as male-specific either if they were significant in males only or if they had significantly stronger effects in males when significant in both sexes. Similarly, those proteins were classified as female-specific if they were significant in females only or if they had significantly stronger effects in females when significant in both sexes.

### Prediction models

The identified proteins were used to build prediction models through logistic regression models males and females separately (sex-stratified data) for female-specific and male-specific proteins separately. Age, sex and *APOE* ε4 genotype were included as covariates in the basic models as reference. Stepwise regression was applied to randomly select sex-specific proteins in the model through function stepAIC in the MASS package separately for males and females. To obtain the specificity and sensitivity of the models, receiver operator characteristic (ROC) curves and areas under the curves (AUC) were completed using R package pROC. The roc.test function based on DeLong’s test was conducted to compare AUC values for two models by using the same package. P-values of comparing AUC values for two models was calculated through bootstrapping 2000 times.

### Biological pathways

Enrichment for gene ontology (GO) was performed using annotations from Enrichr^37^ (https://maayanlab.cloud/Enrichr/). Enrichr serves as a comprehensive platform that provides curated gene sets and functions as a search engine to accumulate biological knowledge for facilitating further discoveries. Entrez gene symbols encoding the identified proteins were included as inputs to uncover significant enrichment (FDR ≤0.05 and enrichment >1) for functional annotation with human as a species. We considered 6,162 proteins from SOMAscan platform as a background. Multiple biological processes were considered including Panther pathway,^81^ gene ontology, and GWAS catalog.^82^

Significant terms were visualized using REVIGO (http://revigo.irb.hr/). Briefly, REVIGO is a software that takes long lists of gene ontology terms and summarize them in hierarchical orders by grouping similar terms and removing redundant terms.^38^ Different visualization options are provided for the gene ontology summary.

### Cell type specificity

Cell-type estimate was performed for the “sex-specific” proteins with cell-type data as background and the “sex-specific” proteins as an input. The cell-type data including gene expression data from human astrocytes, neurons, oligodendrocytes, microglia/macrophages, and endothelial cells to determine the degree of specificity to relevant cell types for the proteins included in the input. The data downloaded contained multiple subtypes of astrocytes; we focused only on human mature astrocytes for our analysis.^39^ For each cell type, the average expression level across individuals for each gene was computed. The total expression level for each gene was then calculated by summing up the averages from each cell type. To determine if a gene is cell-type specific, the contributions of different cell types were compared to the total expression of that gene. Specifically, if the top cell type contributes 1.5 times more expression than the second top cell type, then the gene is considered cell-type specific. In total, 5,750 proteins had corresponding genes that were included in the cell-type expression data. To determine enrichment, Fisher’s exact test was used to compare cell-type difference between female-specific proteins and male-specific proteins.

### Drug targets

Ensemble of Multiple Drug Repositioning Approaches (EMUDRA)^41^ were used to predict potential drug targets for the identified “sex-specific” proteins separately for sex groups. EMUDRA uses Commectivity Map dataset containing of 7,056 transcriptomic data of 1,310 drug perturbations^41^. The principle of EMUDRA is based on literature review indicating that treatment resulting in opposite transcriptomic effects could reverse clinical course of a disease. The package creates an expression weighted cosine method to weight the reference drug-induced expression changes. The method was shown to minimize the influence of the uninformative expression changes. Combining the method to previous methods including weighted signed statistic, nonparametric Kolmogorov-Smirnov, Cosine similarity, Pearson correlation and Spearman correlation to increase accuracy, the package weights each drug on expression changes and then ranks drug orders.^41^ That results in a list of drugs that are predicted to have potential effects on the disease and target genes regulated by drugs.

### Interaction network analyses

The identified “sex-specific” proteins were used to build a functional interaction network using ConsensusPathDB separately for females and males. ConsensusPathDB integrates protein-protein, gene-gene, gene-protein interactions, signaling data, and gene regulatory interactions based on 31 prior published databased to yield interactions and visualize the interactions.^40^ To construct interactions in the network, ConsensusPathDB uses six different methods including CAPPIC, common neighbors, geometric embedding, literature evidence, pathway co-occurrence, and sematic similarity.^40^ Accuracy of the network is estimated by using interaction confidence based on IntScore tool.^40^ High-quality and medium-quality interactions are defined as ≥95% and ≥50%, respectively. In this study, all gene-gene, gene-protein, protein-protein interactions for input proteins were considered. Only medium and high confidence rankings were included in the network to ensure the quality of the network.

## Data Availability

Data availability is upon the request

## Tables

Supplementary Table 1. AD sex-specific analytes stratified by sex in the discovery cohort after meta-analysis

Supplementary Table 2. Concordance of effect sizes in the models stratified by sex between discovery and validation

Supplementary Table 3. List of sex-specific proteins included in prediction models after stepwise regression

Supplementary Table 4. AD sex-specific analytes stratified by gender and APOE ε4 in the discovery cohort

Supplementary Table 5. Gene Ontology of 330 male-specific proteins from Enrichr

Supplementary Table 6. GWAS phenotypes of 330 male-specific proteins from Enrichr

Supplementary Table 7. Gene Ontology of 121 female-specific proteins from Enrichr

Supplementary Table 8. Human pathways of 121 female-specific proteins from Enrichr

Supplementary Table 9. Drug target of 451 sex-specific proteins from EMUDRA

## Acknowledgements

We thank all the participants and their families, as well as the many involved institutions and their staff.

Funding: This work was supported by grants from the National Institutes of Health (R01AG044546 (CC), P01AG003991(CC), RF1AG053303 (CC), RF1AG058501 (CC), U01AG058922 (CC), RF1AG074007 (YJS)), the Michael J. Fox Foundation (CC), the Alzheimer’s Association Zenith Fellows Award (ZEN-22-848604, awarded to CC), and an Anonymous foundation.

The recruitment and clinical characterization of research participants at Washington University were supported by NIH P30AG066444 (JCM), P01AG03991(JCM), and P01AG026276(JCM).

This work was supported by access to equipment made possible by the Hope Center for Neurological Disorders, the Neurogenomics and Informatics Center (NGI: https://neurogenomics.wustl.edu/)and the Departments of Neurology and Psychiatry at Washington University School of Medicine.

<u>ADNI acknowledgement:</u> Data collection and sharing for this project was funded by the Alzheimer’s Disease Neuroimaging Initiative (ADNI) (National Institutes of Health Grant U01 AG024904) and DOD ADNI (Department of Defense award number W81XWH-12-2-0012). ADNI is funded by the National Institute on Aging, the National Institute of Biomedical Imaging and Bioengineering, and through generous contributions from the following: AbbVie, Alzheimer’s Association; Alzheimer’s Drug Discovery Foundation; Araclon Biotech; BioClinica, Inc.; Biogen; Bristol-Myers Squibb Company; CereSpir, Inc.; Cogstate; Eisai Inc.; Elan Pharmaceuticals, Inc.; Eli Lilly and Company; EuroImmun; F. Hoffmann-La Roche Ltd and its affiliated company Genentech, Inc.; Fujirebio; GE Healthcare; IXICO Ltd.; Janssen Alzheimer Immunotherapy Research & Development, LLC.; Johnson & Johnson Pharmaceutical Research & Development LLC.; Lumosity; Lundbeck; Merck & Co., Inc.; Meso Scale Diagnostics, LLC.; NeuroRx Research; Neurotrack Technologies; Novartis Pharmaceuticals Corporation; Pfizer Inc.; Piramal Imaging; Servier; Takeda Pharmaceutical Company; and Transition Therapeutics. The Canadian Institutes of Health Research is providing funds to support ADNI clinical sites in Canada. Private sector contributions are facilitated by the Foundation for the National Institutes of Health (www.fnih.org). The grantee organization is the Northern California Institute for Research and Education, and the study is coordinated by the Alzheimer’s Therapeutic Research Institute at the University of Southern California. ADNI data are disseminated by the Laboratory for Neuro Imaging at the University of Southern California

## References

1. Rajan, K.B., et al. Population estimate of people with clinical Alzheimer’s disease and mild cognitive impairment in the United States (2020-2060). Alzheimers Dement 17, 1966–1975 (2021).

2. Knopman, D.S., et al. Alzheimer disease. Nat Rev Dis Primers 7, 33 (2021).

3. Jessen, F., et al. A conceptual framework for research on subjective cognitive decline in preclinical Alzheimer’s disease. Alzheimers Dement 10, 844–852 (2014).

4. Scheltens, P., et al. Alzheimer’s disease. Lancet 397, 1577–1590 (2021).

5. van Dyck, C.H., et al. Lecanemab in Early Alzheimer’s Disease. N Engl J Med 388, 9–21 (2023).

6. Hebert, L.E., Weuve, J., Scherr, P.A. & Evans, D.A. Alzheimer disease in the United States (2010-2050) estimated using the 2010 census. Neurology 80, 1778–1783 (2013).

7. Collaborators, G.B.D.N. Global, regional, and national burden of neurological disorders, 1990-2016: a systematic analysis for the Global Burden of Disease Study 2016. Lancet Neurol 18, 459-480 (2019).

8. Del-Aguila, J.L., et al. Assessment of the Genetic Architecture of Alzheimer’s Disease Risk in Rate of Memory Decline. J Alzheimers Dis 62, 745–756 (2018).

9. Guo, L., Zhong, M.B., Zhang, L., Zhang, B. & Cai, D. Sex Differences in Alzheimer’s Disease: Insights From the Multiomics Landscape. Biol Psychiatry 91, 61–71 (2022).

10. Deming, Y., et al. Sex-specific genetic predictors of Alzheimer’s disease biomarkers. Acta Neuropathol 136, 857–872 (2018).

11. Aguirre, C.C. & Baudry, M. Progesterone reverses 17beta-estradiol-mediated neuroprotection and BDNF induction in cultured hippocampal slices. Eur J Neurosci 29, 447–454 (2009).

12. Fisher, D.W., Bennett, D.A. & Dong, H. Sexual dimorphism in predisposition to Alzheimer’s disease. Neurobiol Aging 70, 308–324 (2018).

13. Fukumoto, N., et al. Sexually dimorphic effect of the Val66Met polymorphism of BDNF on susceptibility to Alzheimer’s disease: New data and meta-analysis. Am J Med Genet B Neuropsychiatr Genet 153B, 235–242 (2010).

14. Eissman, J.M., et al. Sex differences in the genetic architecture of cognitive resilience to Alzheimer’s disease. Brain 145, 2541–2554 (2022).

15. Aberg, D., et al. Increased Cerebrospinal Fluid Level of Insulin-like Growth Factor-II in Male Patients with Alzheimer’s Disease. J Alzheimers Dis 48, 637–646 (2015).

16. Piccio, L., et al. Cerebrospinal fluid soluble TREM2 is higher in Alzheimer disease and associated with mutation status. Acta Neuropathol 131, 925–933 (2016).

17. Viswanathan, J., et al. An association study between granulin gene polymorphisms and Alzheimer’s disease in Finnish population. Am J Med Genet B Neuropsychiatr Genet 150B, 747–750 (2009).

18. Sun, L.L., Yang, S.L., Sun, H., Li, W.D. & Duan, S.R. Molecular differences in Alzheimer’s disease between male and female patients determined by integrative network analysis. J Cell Mol Med 23, 47–58 (2019).

19. Winkler, J.M. & Fox, H.S. Transcriptome meta-analysis reveals a central role for sex steroids in the degeneration of hippocampal neurons in Alzheimer’s disease. BMC Syst Biol 7, 51 (2013).

20. Paranjpe, M.D., et al. Sex-Specific Cross Tissue Meta-Analysis Identifies Immune Dysregulation in Women With Alzheimer’s Disease. Front Aging Neurosci 13, 735611 (2021).

21. Brooks, L.R.K. & Mias, G.I. Data-Driven Analysis of Age, Sex, and Tissue Effects on Gene Expression Variability in Alzheimer’s Disease. Front Neurosci 13, 392 (2019).

22. Sanfilippo, C., et al. Sex difference in CHI3L1 expression levels in human brain aging and in Alzheimer’s disease. Brain Res 1720, 146305 (2019).

23. Santiago, J.A., Quinn, J.P. & Potashkin, J.A. Sex-specific transcriptional rewiring in the brain of Alzheimer’s disease patients. Front Aging Neurosci 14, 1009368 (2022).

24. Bonham, L.W., et al. CXCR4 involvement in neurodegenerative diseases. Transl Psychiatry 8, 73 (2018).

25. Cao, M., Li, H., Zhao, J., Cui, J. & Hu, G. Identification of age- and gender-associated long noncoding RNAs in the human brain with Alzheimer’s disease. Neurobiol Aging 81, 116–126 (2019).

26. Reumiller, C.M., et al. Gender-related increase of tropomyosin-1 abundance in platelets of Alzheimer’s disease and mild cognitive impairment patients. J Proteomics 178, 73–81 (2018).

27. Yang, H., et al. Prognostic polypeptide blood plasma biomarkers of Alzheimer’s disease progression. J Alzheimers Dis 40, 659–666 (2014).

28. Mahady, L., et al. HDAC2 dysregulation in the nucleus basalis of Meynert during the progression of Alzheimer’s disease. Neuropathol Appl Neurobiol 45, 380–397 (2019).

29. Maffioli, E., et al. Insulin and serine metabolism as sex-specific hallmarks of Alzheimer’s disease in the human hippocampus. Cell Rep 40, 111271 (2022).

30. Wingo, A.P., et al. Sex differences in brain protein expression and disease. Nat Med 29, 2224–2232 (2023).

31. Jack, C.R., Jr., et al. NIA-AA Research Framework: Toward a biological definition of Alzheimer’s disease. Alzheimers Dement 14, 535–562 (2018).

32. Timsina, J., et al. Harmonization of CSF and imaging biomarkers for Alzheimer’s disease biomarkers: need and practical applications for genetics studies and preclinical classification. bioRxiv (2023).

33. Altmann, A., Tian, L., Henderson, V.W., Greicius, M.D. & Alzheimer’s Disease Neuroimaging Initiative, I. Sex modifies the APOE-related risk of developing Alzheimer disease. Ann Neurol 75, 563–573 (2014).

34. Neu, S.C., et al. Apolipoprotein E Genotype and Sex Risk Factors for Alzheimer Disease: A Meta-analysis. JAMA Neurol 74, 1178–1189 (2017).

35. Babapour Mofrad, R., et al. Sex differences in CSF biomarkers vary by Alzheimer disease stage and APOE epsilon4 genotype. Neurology 95, e2378–e2388 (2020).

36. Wang, Y.T., et al. Interactive rather than independent effect of APOE and sex potentiates tau deposition in women. Brain Commun 3, fcab126 (2021).

37. Kuleshov, M.V., et al. Enrichr: a comprehensive gene set enrichment analysis web server 2016 update. Nucleic Acids Res 44, W90–97 (2016).

38. Supek, F., Bosnjak, M., Skunca, N. & Smuc, T. REVIGO summarizes and visualizes long lists of gene ontology terms. PLoS One 6, e21800 (2011).

39. Zhang, Y., et al. Purification and Characterization of Progenitor and Mature Human Astrocytes Reveals Transcriptional and Functional Differences with Mouse. Neuron 89, 37–53 (2016).

40. Herwig, R., Hardt, C., Lienhard, M. & Kamburov, A. Analyzing and interpreting genome data at the network level with ConsensusPathDB. Nat Protoc 11, 1889–1907 (2016).

41. Zhou, X., Wang, M., Katsyv, I., Irie, H. & Zhang, B. EMUDRA: Ensemble of Multiple Drug Repositioning Approaches to improve prediction accuracy. Bioinformatics 34, 3151–3159 (2018).

42. Scott, L.J. & Goa, K.L. Verteporfin. Drugs Aging 16, 139–146; discussion 147-138 (2000).

43. Eshraghi, M., et al. Enhancing autophagy in Alzheimer’s disease through drug repositioning. Pharmacol Ther 237, 108171 (2022).

44. Sumsuzzman, D.M., Choi, J., Jin, Y. & Hong, Y. Neurocognitive effects of melatonin treatment in healthy adults and individuals with Alzheimer’s disease and insomnia: A systematic review and meta-analysis of randomized controlled trials. Neurosci Biobehav Rev 127, 459–473 (2021).

45. Benito-Leon, J., Contador, I., Vega, S., Villarejo-Galende, A. & Bermejo-Pareja, F. Non-steroidal anti-inflammatory drugs use in older adults decreases risk of Alzheimer’s disease mortality. PLoS One 14, e0222505 (2019).

46. Rivers-Auty, J., Mather, A.E., Peters, R., Lawrence, C.B. & Brough, D. Anti-inflammatories in Alzheimer’s disease-potential therapy or spurious correlate? Brain Commun 2, fcaa109 (2020).

47. Borghans, L., Sambeth, A. & Blokland, A. Biperiden Selectively Impairs Verbal Episodic Memory in a Dose- and Time-Dependent Manner in Healthy Subjects. J Clin Psychopharmacol 40, 30–37 (2020).

48. Lee, S.S., et al. Inhibitory effects of ethaverine, a homologue of papaverine, on monoamine oxidase activity in mouse brain. Biol Pharm Bull 24, 838–840 (2001).

49. Oswald, W.J. & Baeder, D.H. Pharmacology of ethaverine HC1: human and animal studies. South Med J 68, 1481–1484 (1975).

50. Lopez-Arrieta, J.M. & Birks, J. Nimodipine for primary degenerative, mixed and vascular dementia. Cochrane Database Syst Rev, CD000147 (2002).

51. Zink, C.F., et al. Nimodipine improves cortical efficiency during working memory in healthy subjects. Transl Psychiatry 10, 372 (2020).

52. Kong, L., Zhang, X., Li, C. & Zhou, L. Potential therapeutic targets and small molecular drugs for pediatric B-precursor acute lymphoblastic leukemia treatment based on microarray data. Oncol Lett 14, 1543–1549 (2017).

53. Simren, J., et al. The diagnostic and prognostic capabilities of plasma biomarkers in Alzheimer’s disease. Alzheimers Dement 17, 1145–1156 (2021).

54. Mila-Aloma, M., et al. Plasma p-tau231 and p-tau217 as state markers of amyloid-beta pathology in preclinical Alzheimer’s disease. Nat Med 28, 1797–1801 (2022).

55. Kac, P.R., et al. Diagnostic value of serum versus plasma phospho-tau for Alzheimer’s disease. Alzheimers Res Ther 14, 65 (2022).

56. Altomare, D., et al. Plasma biomarkers for Alzheimer’s disease: a field-test in a memory clinic. J Neurol Neurosurg Psychiatry 94, 420–427 (2023).

57. Chakroborty, S., et al. Early presynaptic and postsynaptic calcium signaling abnormalities mask underlying synaptic depression in presymptomatic Alzheimer’s disease mice. J Neurosci 32, 8341–8353 (2012).

58. Chang, T.Y., Yamauchi, Y., Hasan, M.T. & Chang, C. Cellular cholesterol homeostasis and Alzheimer’s disease. J Lipid Res 58, 2239–2254 (2017).

59. Lan, G., et al. Presynaptic membrane protein dysfunction occurs prior to neurodegeneration and predicts faster cognitive decline. Alzheimers Dement (2022).

60. Green, K.N., Billings, L.M., Roozendaal, B., McGaugh, J.L. & LaFerla, F.M. Glucocorticoids increase amyloid-beta and tau pathology in a mouse model of Alzheimer’s disease. J Neurosci 26, 9047–9056 (2006).

61. Lesuis, S.L., Weggen, S., Baches, S., Lucassen, P.J. & Krugers, H.J. Targeting glucocorticoid receptors prevents the effects of early life stress on amyloid pathology and cognitive performance in APP/PS1 mice. Transl Psychiatry 8, 53 (2018).

62. Chen, X., et al. Microglia-mediated T cell infiltration drives neurodegeneration in tauopathy. Nature 615, 668–677 (2023).

63. Zhang, L., et al. Sex-specific DNA methylation differences in Alzheimer’s disease pathology. Acta Neuropathol Commun 9, 77 (2021).

64. Guo, L., et al. Sex specific molecular networks and key drivers of Alzheimer’s disease. Mol Neurodegener 18, 39 (2023).

65. Knafo, S., et al. PTEN recruitment controls synaptic and cognitive function in Alzheimer’s models. Nat Neurosci 19, 443–453 (2016).

66. Knafo, S. & Esteban, J.A. PTEN: Local and Global Modulation of Neuronal Function in Health and Disease. Trends Neurosci 40, 83–91 (2017).

67. Sanchez-Puelles, C., et al. PTEN Activity Defines an Axis for Plasticity at Cortico-Amygdala Synapses and Influences Social Behavior. Cereb Cortex 30, 505–524 (2020).

68. Ji, Z., et al. The forkhead transcription factor FOXK2 promotes AP-1-mediated transcriptional regulation. Mol Cell Biol 32, 385–398 (2012).

69. Scopa, C., et al. JUN upregulation drives aberrant transposable element mobilization, associated innate immune response, and impaired neurogenesis in Alzheimer’s disease. Nat Commun 14, 8021 (2023).

70. Ding, P., Gorenflo, M.P., Zhu, X. & Xu, R. Aspirin Use and Risk of Alzheimer’s Disease: A 2-Sample Mendelian Randomization Study. J Alzheimers Dis 92, 989–1000 (2023).

71. Roy, J., et al. Role of melatonin in Alzheimer’s disease: From preclinical studies to novel melatonin-based therapies. Front Neuroendocrinol 65, 100986 (2022).

72. Sanz, J.M., et al. Nimodipine inhibits IL-1beta release stimulated by amyloid beta from microglia. Br J Pharmacol 167, 1702–1711 (2012).

73. Sambeth, A., Riedel, W.J., Klinkenberg, I., Kahkonen, S. & Blokland, A. Biperiden selectively induces memory impairment in healthy volunteers: no interaction with citalopram. Psychopharmacology (Berl) 232, 1887–1897 (2015).

74. Bakker, C., et al. Biperiden Challenge Model in Healthy Elderly as Proof-of-Pharmacology Tool: A Randomized, Placebo-Controlled Trial. J Clin Pharmacol 61, 1466–1478 (2021).

75. Modeste, E.S., et al. Quantitative proteomics of cerebrospinal fluid from African Americans and Caucasians reveals shared and divergent changes in Alzheimer’s disease. Mol Neurodegener 18, 48 (2023).

76. Gold, L., et al. Aptamer-based multiplexed proteomic technology for biomarker discovery. PLoS One 5, e15004 (2010).

77. Yang, C., et al. Genomic atlas of the proteome from brain, CSF and plasma prioritizes proteins implicated in neurological disorders. Nat Neurosci 24, 1302–1312 (2021).

78. Timsina, J., et al. Comparative Analysis of Alzheimer’s Disease Cerebrospinal Fluid Biomarkers Measurement by Multiplex SOMAscan Platform and Immunoassay-Based Approach. J Alzheimers Dis 89, 193–207 (2022).

79. Han, B. & Eskin, E. Random-effects model aimed at discovering associations in meta-analysis of genome-wide association studies. Am J Hum Genet 88, 586–598 (2011).

80. Winkler, T.W., et al. EasyStrata: evaluation and visualization of stratified genome-wide association meta-analysis data. Bioinformatics 31, 259–261 (2015).

81. Mi, H. & Thomas, P. PANTHER pathway: an ontology-based pathway database coupled with data analysis tools. Methods Mol Biol 563, 123–140 (2009).

82. Sollis, E., et al. The NHGRI-EBI GWAS Catalog: knowledgebase and deposition resource. Nucleic Acids Res 51, D977–D985 (2023).

